# Point-of-care HIV viral load testing in a community antiretroviral therapy delivery programme: a randomised controlled trial (PHILA)

**DOI:** 10.1101/2025.05.20.25327934

**Authors:** Jienchi Dorward, Kwena Tlhaku, Yukteshwar Sookrajh, Pedzisai Munatsi, Jessica Naidoo, Emelda Tselana, Andile Maphumulo, Nokuthandwa Mbambo, Thobile Mhlongo-Gumbi, Jennifer A Brown, Lara Lewis, Pravikrishnen Moodley, Natasha Samsunder, Paul K. Drain, Christopher C Butler, Gail Hayward, Nigel Garrett

## Abstract

**Background:** Community antiretroviral therapy (ART) delivery programmes allow people living with HIV (PLWH) to collect treatment nearer to home rather than having to attend clinics. However, delays in laboratory based viral load (VL) testing can prevent timely assessment of eligibility for community ART prescription renewals. We aimed to determine if point-of-care VL testing in the clinic could prevent delayed renewal of community ART prescriptions within the Centralised Chronic Medication Dispensing and Distribution programme (CCMDD) in South Africa.

**Methods:** We conducted an open-label, randomised controlled trial of point-of-care VL testing versus standard laboratory-based VL testing among PLWH who needed renewal of their community ART CCMDD prescription in a single clinic in Durban, South Africa. The primary outcome was renewal of community ART CCMDD prescription by three weeks.

**Results:** We enrolled 200 participants between August 15^th^, 2022 and August 24^th^, 2023. Median age was 44 years (interquartile range [IQR] 37-49), and 65.5% were female. In the intervention arm, 93/100 (93.0%) of participants had a community ART CCMDD prescription renewal within three weeks, compared to 81/100 (81.0%) in the standard-of-care arm (risk difference [RD] 12.0%, 95% confidence interval [CI] 2.9 to 21.2%, p=0.021). Participants received their enrolment VL results after a median of 0 days (IQR 0 to 0) in the intervention arm and 20 days (IQR 7 to not received) in the standard-of-care arm. There was no difference between arms in the proportion retained in care between 8 and 16 weeks (89.0% versus 87.0%, RD 2.0% 95% CI -8.0 to 12.0). The mean number of clinic visits required for community ART CCMDD prescription renewal was lower in the intervention arm (1.06) versus the standard-of-care arm (1.60, RD -0.54, 95% CI -0.40 to -0.68), as was the total travel cost to participants to have their CCMDD prescription renewed (South African Rands [ZAR] 47.7 versus ZAR 72.8, RD ZAR -25.1 [95% CI -9.2 to -41.1]).

**Conclusions:** Point-of-care VL testing improved renewal of community ART prescriptions in South Africa, by reducing time to results for healthcare workers and clients, and reducing the number of clinic visits and associated travel costs.

**Trial registration:** Pan-African Clinical Trials Registry: PACTR202002785960123

## INTRODUCTION

South Africa has the largest antiretroviral therapy (ART) programme globally, with over 5 million people receiving ART. In order to provide chronic medication efficiently, including ART, the South African government introduced the Centralised Chronic Medication Dispensing and Distribution system (CCMDD),^1^ which provides ART to over 2 million people.^2^ CCMDD can benefit people living with HIV (PLWH) by facilitating ART collection at community-based pick-up points (churches, community groups, pharmacies) that are nearer, have longer opening hours and are less congested than clinics.^3 4^ However, barriers to implementation of CCMDD^3 5^ have resulted in between 16-26% of CCMDD participants being ‘dormant’,^6^ meaning their CCMDD prescriptions have lapsed by more than three weeks.

Reducing the number of dormant clients has been a South African National Department of Health priority, but has proved difficult. One potential cause is the blood tests required to determine eligibility for CCMDD, as only people with a suppressed annual HIV viral load (VL) <50 copies/mL are eligible for CCMDD. Determining viral suppression can be a barrier to enrolment and continuation in CCMDD, as testing is conducted at centralized laboratories, meaning clients generally have to attend a clinic twice for a blood draw and then review of results to determine CCMDD eligibility, with results sometimes being delayed or lost (Figure S1A).^7^ This can lead to delays in renewal of CCMDD prescriptions, resulting in clients continuing to attend clinics rather than collecting ART in the community.

Point-of-care VL testing may help mitigate this problem and reduce the number of dormant clients, by providing same day results that allow CCMDD referral in one clinic visit.^8^ However, whether point-of-care VL testing can be successfully integrated into CCMDD clinical pathways, with results used by routine, front-line healthcare workers is not known. Therefore, we aimed to assess whether point-of-care VL testing could increase CCMDD renewals and reduce time to ART collection in CCMDD in a public clinic in South Africa.

## METHODS AND ANALYSIS

### Trial design

We conducted the Point-of-care HIV viral Load testing in a community Antiretroviral therapy delivery programme (PHILA) study, a single-site, open-label, individually randomised trial of point-of-care HIV VL testing among PLWH in the CCMDD programme. The full protocol is available online.^9^

The study was conducted at the Prince Cyril Zulu Communicable Disease Centre (PCZ CDC), a large, public clinic next to the central Durban transport hub, with support from the adjacent Centre for the AIDS Programme of Research in South Africa (CAPRISA) eThekwini Clinical Research Site. PCZ CDC provides HIV, tuberculosis and other primary care services and is in the province of KwaZulu-Natal, which has an estimated HIV prevalence of 27% among adults aged 15-49^10^. HIV treatment is provided free at the point of service, according to South African Department of Health guidelines,^11^ which at the time of the study recommended HIV VL testing at six months after ART initiation, and then annually. People were eligible for CCMDD if they were clinically stable with a VL <50 copies/ml and estimated glomerular filtration rate (eGFR) >50 mL/min/1.73 m^2^ in the past 12 months, not pregnant, and did not have any medical condition requiring frequent clinical monitoring such as tuberculosis or uncontrolled diabetes or hypertension. Eligible people could be referred by a clinic nurse, who would prescribe six months of ART, with the first two or three months provided at the referral clinic visit. After two or three months, the client would then collect a further three-month supply of ART, or two further two-month supplies, at the pick-up point of their choice, before the client would return to the clinic for assessment and a new prescription after 6 months (Figure S1A). Every 12 months, the clinic visit assessment would include a VL test, to demonstrate viral suppression. Pick-up points include private pharmacies, community group venues and smart lockers known as Pele-Boxes, from which patients can retrieve pre-packed medication using a pin code sent to their phone.^12^

### Participants

People were eligible for enrolment into PHILA if they were living with HIV, aged ≥18 years, receiving ART in CCMDD and due a VL test at the clinic for a renewal of their CCMDD prescription. People who were known to be pregnant or breast feeding, had current tuberculosis, known diabetes with blood glucose >7.0mmol/L, known hypertension with blood pressure ≥140/90 mmHg, or other medical conditions requiring regular clinical consultations, would not have been eligible to continue in CCMDD and were not eligible for enrolment into PHILA.

### Enrolment and randomisation

After screening and providing informed consent, eligible participants were enrolled and randomised by a study nurse, using a pre-programmed electronic case report form, in a 1:1 ratio to the point-of-care arm or the standard-of-care arm. A statistician generated the allocation sequence using random numbers with variable block sizes. All study staff except the statistician were blinded to the allocation sequence. Clinic staff and participants were told the participant’s allocation at enrolment.

### Interventions

At enrolment and during follow up, clinical management was provided by public-sector clinic counsellors, nurses or clinicians, according to South African Department of Health guidelines.^11^ VL and creatinine testing was conducted according to the study arm.

#### Point-of-care testing

For participants randomised to the point-of-care arm, creatinine testing was conducted by a research nurse from a capillary finger-prick sample using the Statsensor Xpress-I (Nova Biomedical, Waltham, USA), which provides a result in 90 seconds. VL testing was conducted using the Xpert HIV-1 VL XC assay (Cepheid, Sunnyvale, USA), which measures quantitative VL in 90 minutes. We planned for clinic nurses to do the point-of-care VL testing in the study clinic, but staff shortages and COVID-19 related disruption meant that it was initially conducted on a 4 module GeneXpert machine by a research laboratory technician at the clinic site laboratory. Later, research nurse testing in a clinical room using two single module GeneXpert machines was introduced, with support from laboratory technicians. Both laboratory technicians and nurses received on-site manufacturer training and ongoing support to conduct testing according to manufacturer instructions. In brief, for both laboratory technician and nurse testing, a venous blood sample was centrifuged to provide 1ml of plasma, which was tested using the Xpert HIV-1 VL XC cartridge. In the case of invalid results, leftover plasma was used for retesting, or a repeat sample was requested. Participants were encouraged to wait for the result (Figure S1), but if they were not willing, results were provided at their next clinic appointment, scheduled by clinic staff in consultation with participants at the soonest possible date. Once point-of-care results were available, they were provided to routine clinic staff. Those with a VL <50 copies/mL and eGFR >50 mL/min/1.73 m^2^ were referred to CCMDD, while those with a VL ≥50 copies/mL or eGFR ≤50 mL/min/1.73 m^2^ were not eligible for CCMDD and so were referred to a counsellor or clinician for further management.

#### Laboratory-based VL testing

For participants randomised to the standard-of-care arm, venous blood was drawn by a research nurse and sent for creatinine and VL testing off-site by the National Health Laboratory Service. VL testing was conducted using the Alinity m HIV-1 VL analyser (Abbott, Chicago, USA). Initially, participants would return to the clinic after 7 to 28 days to be assessed for CCMDD eligibility based on VL and creatinine results (Figure S1A). However, almost exactly halfway through enrolment, on 24^th^ May 2023, new South African guidelines were implemented which recommended that in the standard-of-care arm, clients could have their CCMDD prescription renewed without waiting for the laboratory results (Figure S1B), and that they would only be called back if the VL was ≥50 copies/mL or eGFR ≤50 mL/min/1.73 m^2^.^13^ Otherwise, they would receive the results when they returned to clinic six months later.

### Follow-up

Participants were not followed up in person by research staff. Instead, outcomes were ascertained up to 16 weeks using reviews of participants’ routine clinical charts, laboratory results and the CCMDD electronic database. If by 16 to 18 weeks there was no evidence of CCMDD renewal prescription or ART collection, the research team attempted to contact the participant to establish the reason for not collecting ART in CCMDD.

### Outcomes

The primary outcome was the proportion of participants in each arm with renewal of their CCMDD prescription within three weeks of enrolment. We chose this outcome to align with the programmatic definition of being dormant if a CCMDD prescription had lapsed by three weeks. Secondary outcomes were the number of days from enrolment to CCMDD prescription renewal, days to first CCMDD ART collection, proportion of participants in each arm retained in care (defined as documented ART collection between 6 to 16 weeks post-enrolment), days from enrolment until participants received their VL results, number of clinic visits from enrolment until CCMDD prescription, and travel costs in South African Rands (ZAR, 1 ZAR ∼ 18 United States Dollars) from the patient perspective to have their CCMDD prescription renewed. As per the study protocol, because PHILA did not involve an investigational medicinal product or intervention that affects physiology, only adverse events of hospitalisations and deaths were recorded.

### Sample Size and Statistical methods

Assuming that 75% of participants in the standard-of-care arm would achieve the primary outcome of CCMDD renewal by 3 weeks, and a 15% improvement in the point-of-care arm, we calculated that a sample size of 100 participants per arm would give us 80% power to demonstrate superiority with a two-sided alpha of 0.05%.

In pre-specified analyses, we calculated the proportions of participants achieving study outcomes and compared proportions in each arm using the chi-squared test. We included all participants enrolled in each arm in intention-to-treat analyses. For secondary time-to-event outcomes, we did not use Cox proportional hazards models due to the high proportion of tied events in the point-of-care arm on days 0 and 1, and the violation of the proportional hazards assumption. Instead, as pre-specified in the protocol, we compared proportions achieving the outcome of interest at 2, 4, 8, 12 and 16 weeks post enrolment, using chi-squared tests and Newcombe Wilson 95% confidence intervals. We also drew Kaplan Meier survival curves. Amongst those with CCMDD renewals, we compared the mean number of clinic visits from enrolment until CCMDD prescription renewal, and the mean total travel cost to the participant to attend these clinic visits (number of visits x travel cost per visit), using Students t-Test. In a post-hoc analysis, we used a Poisson regression model with robust standard errors and an interaction term between time period and study arm, to evaluate whether there was a sub-group effect depending on enrolment before or after the guideline changed to allow same-day CCMDD renewal in the standard-of-care arm without receipt of VL results.

Data was collected and managed using REDCap (Research Electronic Data Capture),^14^ and analysed using R v4.1.^15^

### Post hoc evaluation of the impact of same day laboratory viral load testing and CCMDD prescription renewal in public clinics using routine de-identified data

Lastly, we evaluated the broader impact, outside the PHILA trial, of the guideline change to allow clients to have their CCMDD prescription renewed on the same day as the laboratory VL test, without waiting for the results. Specifically, we aimed to assess whether this resulted in people being referred to CCMDD while still viraemic, and whether those with viraemia were managed according to the guidelines which recommend prompt recall to the clinic for enhanced adherence counselling and a repeat viral load after 3 months. We analysed de-identified, routinely collected TIER.Net^16^ CCMDD referral and viral load data from 108 primary care clinics in KwaZulu-Natal, from 24^th^ May (date of guideline change) to 10^th^ September (16 weeks before data cut of 31^st^ December), 2023. We evaluated 1) the number of non-pregnant adults ≥18 years old in CCMDD who had their community ART CCMDD prescription renewed on the same day as the laboratory VL was taken, 2) the proportion of VLs with viraemia >50 copies/mL and ≥1000 copies/mL, and of these, the proportion who had 3) a clinic visit and 4) a repeat VL within 16 weeks (corresponding to the PHILA follow-up time and the recommendation to have a repeat VL within 3 months).

### Ethical approvals

The University of KwaZulu-Natal Biomedical Research Ethics Committee (BREC/00000837/2019 and BE646/17) and the University of Oxford Tropical Research Ethics Committee (OxTREC 64-19) approved the study. PHILA was registered on the Pan African Clinical Trials Registry (PACTR202002785960123) on 12^th^ February 2020. The University of Oxford is the study sponsor.

## RESULTS

### Study population

Between August 15^th^, 2022 and August 24^th^, 2023, we assessed 899 people in CCMDD for eligibility, and enrolled 200 participants (Figure 1). Median age was 44 years (interquartile range [IQR] 37-49), 65.5% were female, and median time on ART was 8.0 years (IQR 6.0-10.9) (Table 1). 83.0% were receiving tenofovir disoproxil fumarate, lamivudine and dolutegravir (TLD), and 14.5% tenofovir disoproxil fumarate, emtricitabine and efavirenz (TEE). Median time since the participant had been first referred to CCMDD was 3.8 years (IQR 1.8 to 6.0), and median time since the last CCMDD referral was 168 days (IQR 161-172). Median monthly income was ZAR 3500 (IQR 350 to 5000). The median time and cost to travel to the clinic and back was 60 minutes (IQR 50 to 90) and ZAR 44 (28 to 50) respectively. The median time and cost to travel to the last used pick-up-point and back was 50 minutes (IQR 30-60) and ZAR 30 (IQR 20-50) respectively.

**Table 1.**
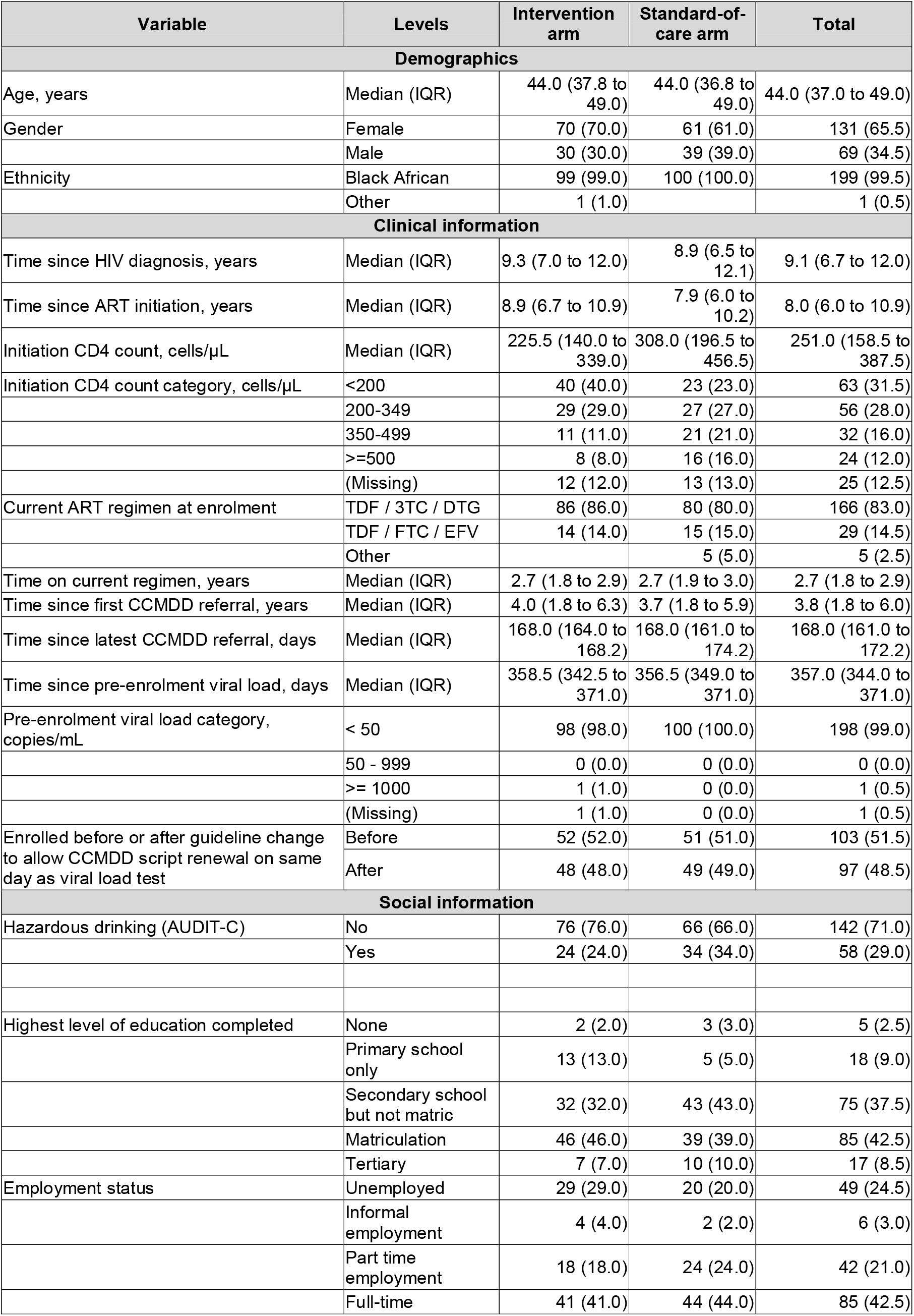
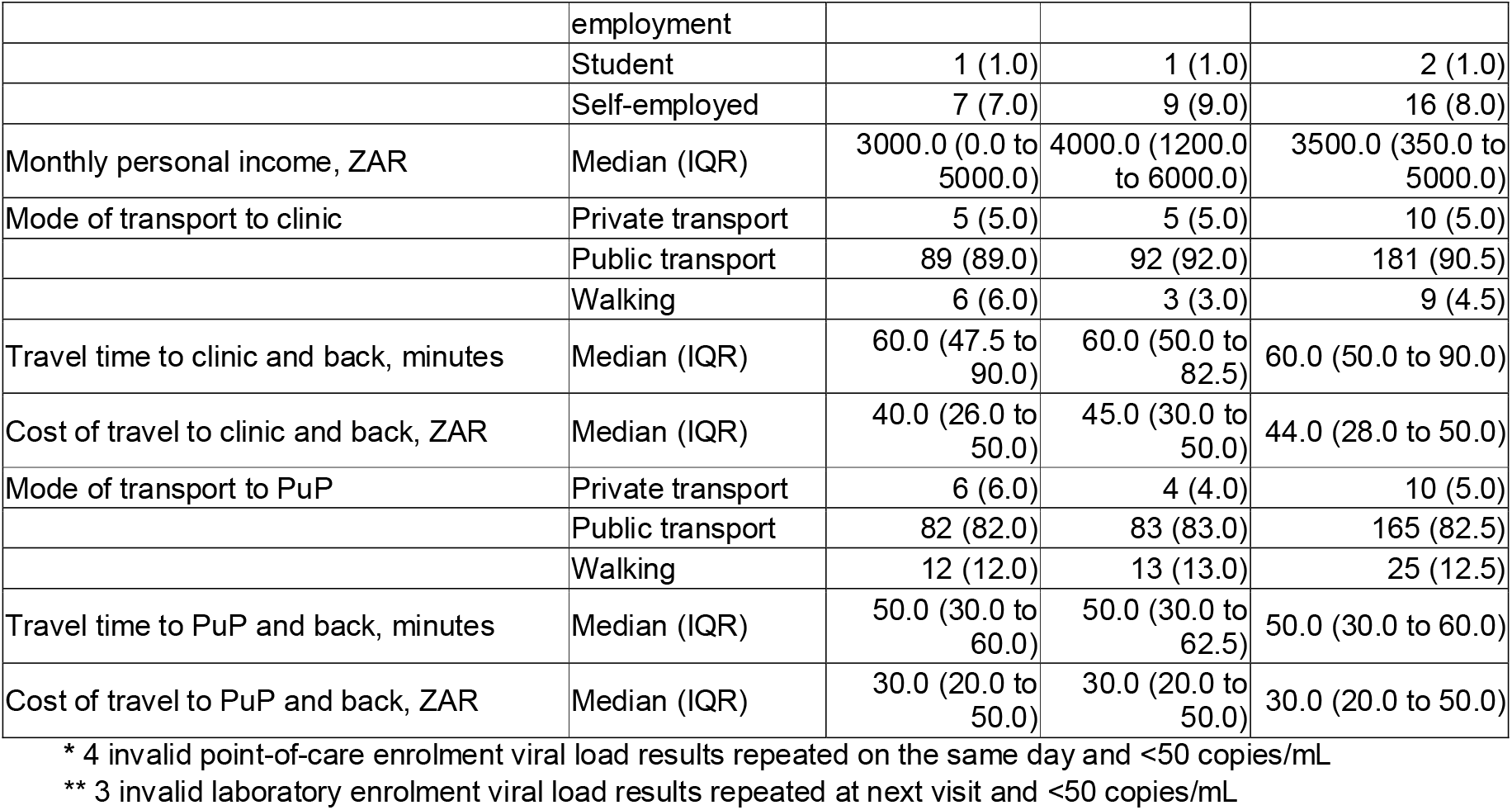
Baseline characteristics of PHILA study participants, n = 200.

**Figure 1.**
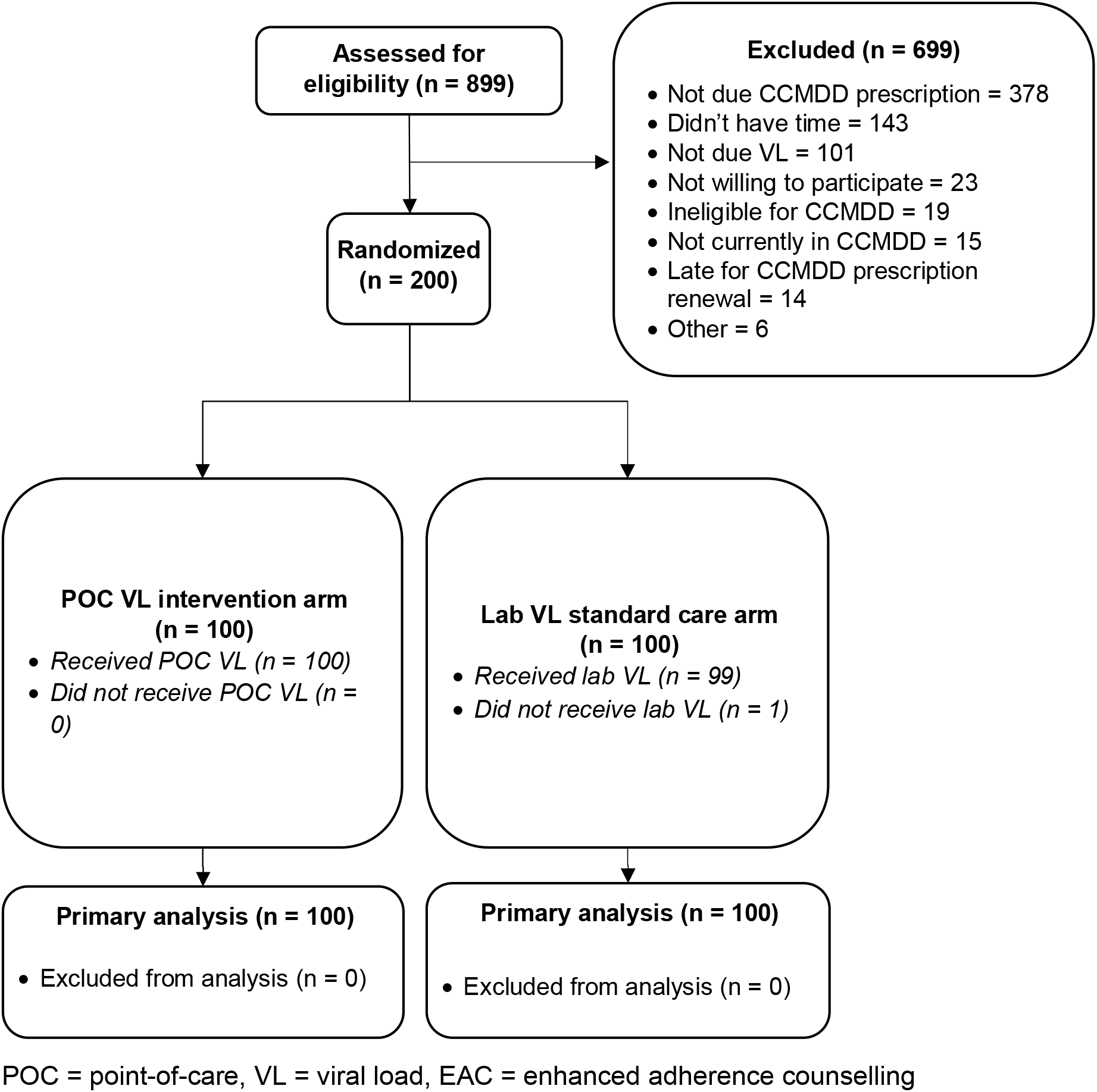
PHILA study CONSORT diagram.

There were slightly higher proportions of women (70% versus 61%) and participants reporting no monthly income (28% versus 19%) in the intervention arm. There were more participants reporting hazardous alcohol use (76% versus 66%) in the standard-of-care arm, and the median CD4 count at initiation (308 versus 226 cells/μL) was higher in the standard-of-care arm. Other demographics and clinic variables were well balanced between the two groups.

Enrolment VLs were suppressed <50 copies/mL for 94% of the intervention arm and 92% of the standard-of-care arm (Table 2). In the intervention arm, site laboratory staff conducted 74 (74%) and nurses 26 (26%) of the point-of-care VL tests. There were four invalid enrolment point-of-care results, which were repeated and results given to participants on the same day; all were <50 copies/mL. There were three invalid enrolment laboratory VL results, which were subsequently repeated after 5, 8 and 45 days from enrolment, with results reaching the patient 99, 11 and 48 days from enrolment respectively.

**Table 2.**
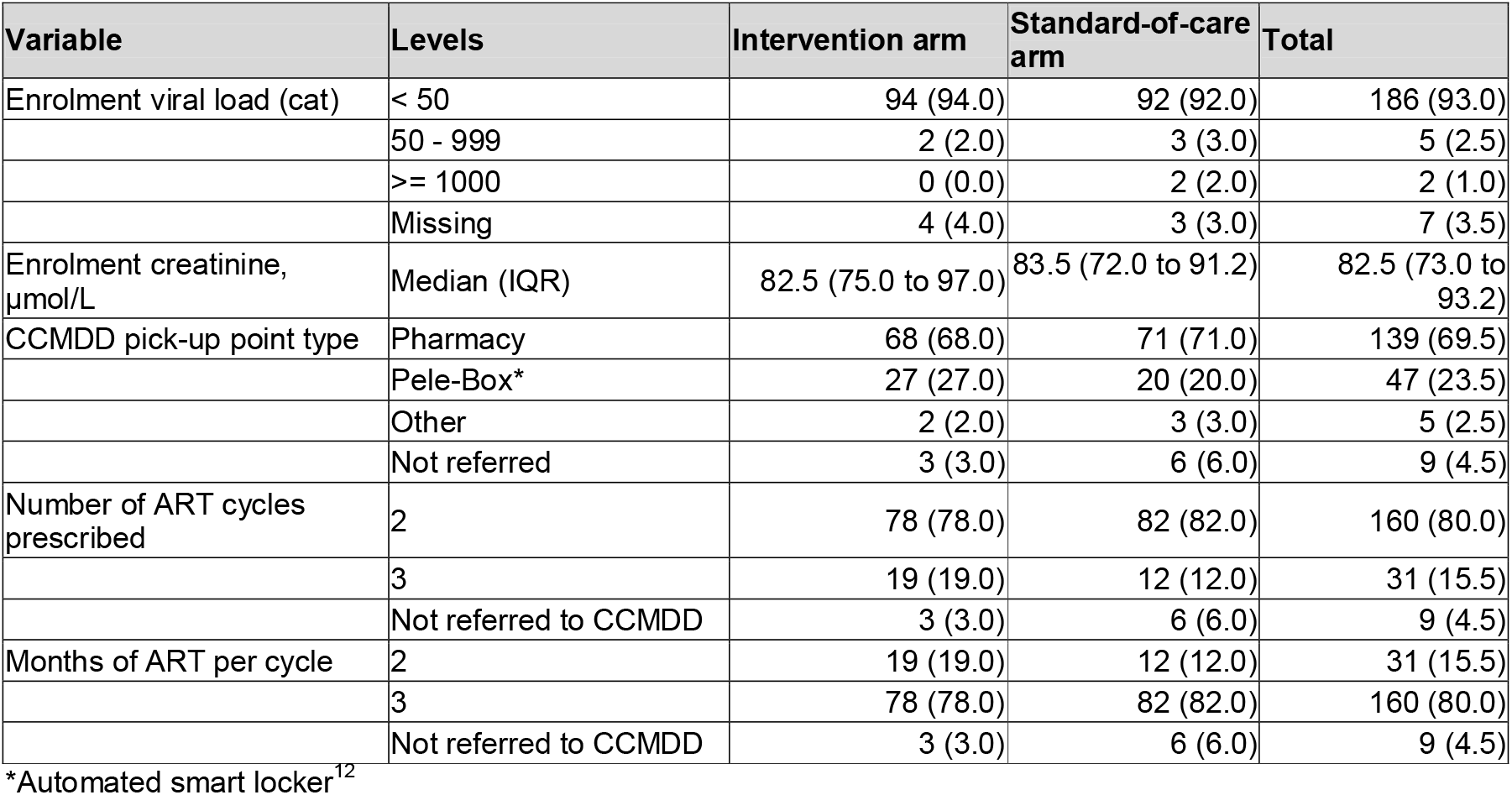
PHILA study enrolment visit viral loads and CCMDD referral data.

| Variable | Levels | Intervention arm | Standard-of-care arm | Total |
| --- | --- | --- | --- | --- |
| Enrolment viral load (cat) | < 50 | 94 (94.0) | 92 (92.0) | 186 (93.0) |
|  | 50 - 999 | 2 (2.0) | 3 (3.0) | 5 (2.5) |
|  | >= 1000 | 0 (0.0) | 2 (2.0) | 2 (1.0) |
|  | Missing | 4 (4.0) | 3 (3.0) | 7 (3.5) |
| Enrolment creatinine, μmol/L | Median (IQR) | 82.5 (75.0 to 97.0) | 83.5 (72.0 to 91.2) | 82.5 (73.0 to 93.2) |
| CCMDD pick-up point type | Pharmacy | 68 (68.0) | 71 (71.0) | 139 (69.5) |
|  | Pele-Box* | 27 (27.0) | 20 (20.0) | 47 (23.5) |
|  | Other | 2 (2.0) | 3 (3.0) | 5 (2.5) |
|  | Not referred | 3 (3.0) | 6 (6.0) | 9 (4.5) |
| Number of ART cycles prescribed | 2 | 78 (78.0) | 82 (82.0) | 160 (80.0) |
|  | 3 | 19 (19.0) | 12 (12.0) | 31 (15.5) |
|  | Not referred to CCMDD | 3 (3.0) | 6 (6.0) | 9 (4.5) |
| Months of ART per cycle | 2 | 19 (19.0) | 12 (12.0) | 31 (15.5) |
|  | 3 | 78 (78.0) | 82 (82.0) | 160 (80.0) |
|  | Not referred to CCMDD | 3 (3.0) | 6 (6.0) | 9 (4.5) |

\*Automated smart locker<sup>12</sup>

### Primary outcome

In the intervention arm, 93/100 (93.0%) of participants had a CCMDD prescription renewal within three weeks, compared to 81/100 (81.0%) in the standard-of-care arm (risk difference [RD] 12.0%, 95% confidence interval [CI] 2.9 to 21.2%, p=0.021, Table 3). In a post-hoc sensitivity analysis, the point estimate of the effect of the intervention was greater prior to the guideline change to allow same day CCMDD prescription renewal (RD 23.6%, 95% CI 10.3 to 36.9), than after (RD 0.2%, 95% CI -12.3 to 11.9), although the likelihood ratio test for an interaction was not significant (p=0.351).

**Table 3.** Primary and secondary outcomes for the PHILA study, n = 200.

|  | Intervention<br>n/N (%) | Standard care<br>n/N (%) | Risk difference<br>(95% CI) | p value |
| --- | --- | --- | --- | --- |
| **Primary outcome** |  |  |  |  |
| CCMDD prescription renewal within 3 weeks | 93/100 (93.0%) | 81/100 (81.0%) | 12.0% (2.9 to 21.2) | 0.021 |
| Before guideline change* | 50/52 (96.2%) | 37/51 (72.5%) | 23.6% (10.3 to 36.9) | 0.351** |
| After guideline change* | 43/48 (89.6%) | 44/49 (89.8%) | -0.2% (-12.3 to 11.9) |  |
| **Secondary outcomes** |  |  |  |  |
| Time to CCMDD prescription renewal, days (median, IQR) | 0 (0, 0) | 7 (0, 12) | - | - |
| Before guideline change | 0 (0, 0) | 7 (8, 21) | - | - |
| After guideline change | 0 (0, 0) | 0 (0, 6) | - | - |
| CCMDD prescription renewal within: |  |  |  |  |
| 2 weeks | 93/100 (93%) | 76/100 (76%) | 17% [6.25 - 27.75] | 0.002 |
| 4 weeks | 93/100 (93%) | 88/100 (88%) | 5% [-4.1 - 14.1] | 0.335 |
| 8 weeks | 95/100 (95%) | 92/100 (92%) | 3% [-4.82 - 10.82] | 0.566 |
| 12 weeks | 95/100 (95%) | 92/100 (92%) | 3% [-4.82 - 10.82] | 0.566 |
| 16 weeks | 97/100 (97%) | 94/100 (94%) | 3% [-3.73 - 9.73] | 0.495 |
| Time to first CCMDD ART collection, days (median, IQR) | 84 (83 to 89) | 91 (84 to 105) | - | - |
| First CCMDD ART collection within: |  |  |  |  |
| 2 weeks | 0/100 (0.0%) | 0/100 (0%) | 0% [0 - 0] | NA |
| 4 weeks | 0/100 (0.0%) | 0/100 (0%) | 0% [0 - 0] | NA |
| 8 weeks | 3/100 (3.0%) | 2/100 (2%) | 1% [-4.33 - 6.33] | 1.000 |
| 12 weeks | 29/100 (29.0%) | 15/100 (15%) | 14% [1.68 - 26.32] | 0.026 |
| 16 weeks | 80/100 (80.0%) | 78/100 (78.0%) | 2.0% [-10.29 - 14.29] | 0.862 |
| Proportion of participants retained in care | 89/100 (89.0%) | 87/100 (87.0%) | 2.0% (-8.0 to 12.0) | 0.828 |
| Time to participant receiving enrolment VL result, days (median, IQR) | 0 (0, 0) | 20 (7, Not received) | - | - |
| Before guideline change | 0 (0, 0) | 8 (7, 21) | - | - |
| After guideline change | 0 (0, 0) | Not received (19, Not received) | - | - |
| Participants who received viral load results within: |  |  |  |  |
| 2 weeks | 100/100 (100%) | 42/100 (42%) | 58% [47.33 - 68.67] | <0.001 |
| 4 weeks | 100/100 (100%) | 59/100 (59%) | 41% [30.36 - 51.64] | <0.001 |
| 8 weeks | 100/100 (100%) | 63/100 (63%) | 37% [26.54 - 47.46] | <0.001 |
| 12 weeks | 100/100 (100%) | 65/100 (65%) | 35% [24.65 - 45.35] | <0.001 |
| 16 weeks | 100/100 (100%) | 65/100 (65%) | 35% [24.65 - 45.35] | <0.001 |
|  | **Mean (95% CI)** | **Mean (95% CI)** | **Mean difference (95% CI)** |  |
| Number of clinic visits required for CCMDD renewal | 1.06 (1.00 - 1.12) | 1.60 (1.47 - 1.73) | -0.54 (-0.40 to -0.68) | <0.001 |
| Travel costs from the patient perspective to have CCMDD prescription renewed (ZAR) | 47.7 (39.6 – 55.8) | 72.8 (59.0 – 86.7) | -25.1 (-9.2 to -41.1) | <0.001 |

\* On May 24th, 2023 new South African guidelines were implemented which recommended that in the standard-of-care arm, clients could have their CCMDD prescription renewed on the same day as their VL was taken, without waiting for the results
\*\*p for interaction using likelihood ratio test
\*\*\*Note the proportional hazards assumption violated

### Secondary outcomes

The median time to CCMDD prescription renewal was 0 days in the intervention arm (IQR 0 to 0) and 7 days in the standard-of-care arm (IQR 0 to 8; Fig 2A). The proportion with CCMDD renewal by 2 weeks was higher in the intervention arm (93/100 (93.0%) versus 76/100 (76.0%), RD 17.0% (6.3 – 27.8) p=0.001), but was not significantly different by 4 weeks onwards (Table 3). By the end of follow-up, 97% in the intervention arm and 94% in the standard-of-care arm had a CCMDD prescription renewal. The most common type of pickup points that participants were referred to were private pharmacies (n=139), followed by ‘Pele Boxes’ (n=47, Table 2). The majority (n=160, 83.8%) were prescribed two cycles of three months of ART, as opposed to three cycles of two months ART supply (n=31, 16.2%), which was more common early in the trial (Figure S2).

**Figure 2.**
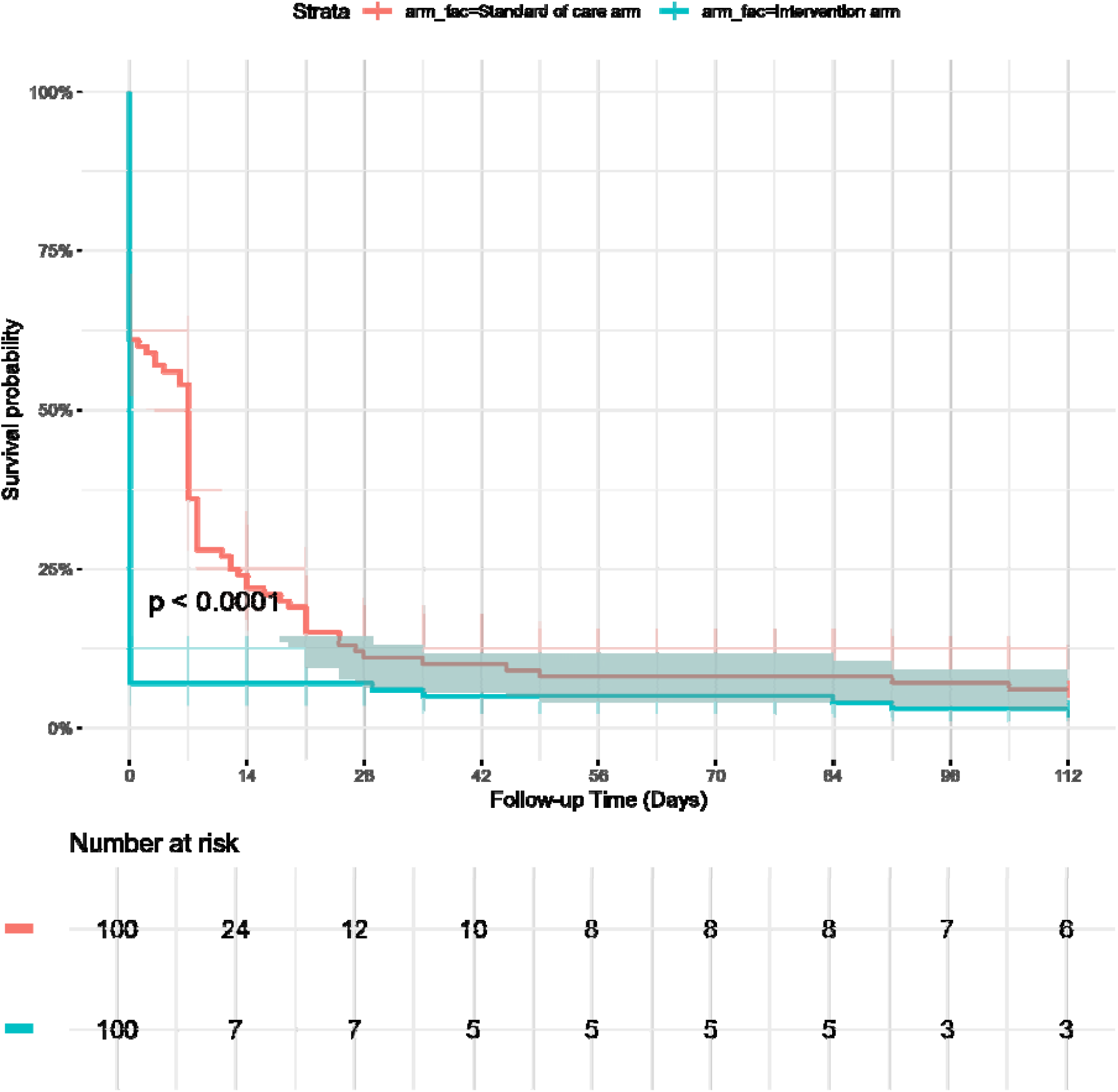
Time to CCMDD prescription renewal in the PHILA trial.

The median time to first CCMDD ART collection was 84 days (IQR 83 to 89) in the intervention arm and 91 (IQR 84 to 105) in the standard-of-care arm (Figure S3). The proportion with CCMDD ART collection at 12 weeks was higher in the intervention arm (29.0%) than in the standard-of-care arm (15%, RD 14.0%, 95% CI 1.7 to 26.3), but not different at any other timepoint. There was no difference between arms in the proportion retained in care between 8 and 16 weeks (89.0% versus 87.0%, RD 2.0% 95% CI -8.0 to 12.0). The mean number of clinic visits required for CCMDD renewal was lower in the intervention arm (1.06) versus the standard-of-care arm (1.60, mean difference -0.54, 95% CI -0.40 to -0.68), as was the total travel cost to participants to have their CCMDD prescription renewed (ZAR 47.7 versus ZAR 72.8, mean difference ZAR -25.1 [95% CI -9.2 to -41.1]).

Participants received their enrolment VL results after a median of 0 days (IQR 0 to 0) in the intervention arm and 20 days (IQR 7 to not received) in the standard-of-care arm (Figure S4), with more participants receiving their results in the intervention versus standard-of-care arm at all timepoints through to the end of follow-up at 16 weeks. Of note, after the guideline change, 34/49 (69.4%) in the standard-of-care arm had their CCMDD prescription renewed on the same day that their enrolment laboratory VL blood was drawn. Of these, 32/34 (94.1%) were <50 copies/mL and 30/32 (93.8%) reached the end of follow up without receiving their VL result. 1/34 (2.9%) had an invalid enrolment laboratory VL and was called back for a repeat test after 5 days, and 1/34 (2.9%) had a VL of 120 copies/mL, but had no further clinic visits before the end of follow-up.

During follow-up, there was one hospitalisation, resulting in death due to pneumonia in the intervention arm. This was not deemed to be related to the intervention.

### Outcomes of CCMDD referral and laboratory VL testing in KwaZulu-Natal clinics

Between 24^th^ May 2023 and 10^th^ September 2023 there were 16,568 adults in CCMDD with a CCMDD prescription renewal on the same day as blood draw in 108 public KwaZulu-Natal clinics (Table S1). Of these, 2,932/16,568 (17.7%) were >50 copies/mL (with 373/16,568, 2.3% ≥1000 copies/mL). Of those with a viral load >50 copies/mL, 25/2932 (0.9%) had a clinical visit with a nurse/doctor within 16 weeks, at a median of 88 days (IQR 84-101), and 111/2,932 (3.8%) had a repeat VL taken within 16 weeks, after a median of 86 (IQR 75.5-94.5) days. Of those with a viral load ≥1000 copies/mL, 2/373 (0.5%) had a clinical visit within 16 weeks (after 96 and 107 days) and 21/373 (5.6%) had a repeat VL within 16 weeks (after a median of 84 (IQR 68-92) days).

## DISCUSSION

### Summary

In this randomised controlled trial we found that point-of-care testing reduced the number of people with a delayed CCMDD prescription renewal of three or more weeks, who are defined as ‘dormant clients’ in the CCMDD programme. Point-of-care testing also reduced the time to receipt of VL results, and the number of clinic visits, and associated travel costs, required by participants for CCMDD prescription renewal.

### Interpretation and comparison with other studies

These results suggest that point-of-care testing is a potential strategy to improve efficiency in the CCMDD programme, and other community ART delivery programmes, by reducing the number of clinic visits (and associated travel costs) required to confirm viral suppression and CCMDD eligibility. However, the change in South African guidelines to allow CCMDD prescription renewal without review of laboratory results also led to quicker renewals and a reduction in clinic visits in the standard-of-care arm, but with the effect of greatly increasing the time to patients receiving their VL results. It is important that people know their VL results to increase self-efficacy and self-management, which has been associated with better engagement in care.^17^ In particular, given that people with an undetectable VL cannot transmit HIV (promoted in the undetectable = untransmittable [U=U] campaign), knowledge of VL may guide sexual behaviour and is important in preventing HIV transmission.^18 19^ The one participant in PHILA who was referred into CCMDD and subsequently turned out to have an unsuppressed VL was not seen again during follow-up. And in our post-hoc analysis in 108 public sector clinics during the early stages of the guideline change, the vast majority of people with viraemia who had their CCMDD prescriptions on the same day, had no follow-up clinic visits or repeat VL within 16 weeks. This shows that systems to review results when clients are not in the clinic need to be improved. Unless these people were phoned (which is not captured in the routine data system) they could have been viraemic, without knowing, until their next clinic visit after 6 months. Models of differentiated service delivery aim to increase efficiency in the healthcare system and also better serve the needs of PLWH, and so balancing the need for efficient CCMDD prescription renewal with provision of results is important. Point-of-care VL testing may help achieve both aims, but additional strategies to provide all VL results, without clients needing to attend clinic should be explored.

Evidence around the impact of point-of-care VL testing varies depending on the type of study. Similar to our findings, several randomised trials have shown the point-of-care VL testing can improve availability of VL results, but none that we are aware of have assessed the impact in a differentiated ART delivery programmes such as those used in CCMDD.^8^ A randomised controlled trial in adults with HIV who had recently initiated ART found point-of-care VL testing was associated with better 12-month viral suppression and retention in care,^20^ but other trials in adults,^21^ children,^22^ pregnant women,^23^ adolescents^24^ and people with viraemia,^25^ have not found an impact on viral suppression and retention outcomes. We did not assess viral suppression, but found no effect on short term retention-in-care. Studies in more routine settings have found more rapid availability of results, but results have not always led to rapid clinical action or improved outcomes.^26-28^ Qualitative research suggests that clients appreciate quicker, same-day VL results, which may facilitate better adherence, but there have been concerns around feasibility of implementing point-of-care VL testing in routine healthcare settings.^29 30^

### Strengths and limitations

Strengths of our study include the randomized design, the high fidelity to the point-of-care VL intervention and the use of routine healthcare staff and settings, with management following South African guidelines. We did not assess a viral suppression outcome, as given the high proportion of people with viral suppression at baseline, it is unlikely that point-of-care VL could improve this outcome. Instead, we focussed on a programmatic outcome measure (dormancy) that is used by the South African Department of Health to measure efficiency in the CCMDD programme, making our results directly relevant to local policy makers. We had planned for VL testing to be conducted by nurses in the clinic, but staff shortages and changes in services from the COVID-19 pandemic meant that research site laboratory staff conducted most of the testing.

### Implications for research and policy

We show that point-of-care VL has potential to improve the efficiency of differentiated ART delivery programmes, without compromising on providing VL results to clients. In differentiated care programs where annual ART prescriptions are used,^31^ point-of-care VL could allow stable clients to attend clinic only once a year.^8^ However, further work to determine what is required to successfully implement point-of-care VL testing in routine, non-research settings is required, and we will present qualitative work around the implementation of point-of-care VL testing in a separate manuscript. Furthermore, better VL result review processes and call back of people with a high VL are required within the current policy of allowing CCMDD renewal on the same day as laboratory VL testing. Alternative strategies to provide VL results remotely, using phone calls or text messaging, should also be explored. While these strategies have been used successfully in high-income settings to provide sexually transmitted infection results,^32^ maintaining confidentiality in low- and middle-income country settings can be difficult due to phones often being shared,^30^ and so further work to develop these strategies is required.^33^

## Conclusion

In this randomized controlled trial we showed that point-of-care VL testing improves the proportion of people with CCMDD ART prescription renewal within three weeks, by reducing time to availability of VL results for healthcare workers and clients, and reduces clinic visits and associated travel costs for clients.

## Data Availability

Bona fide researchers will be able to request access to anonymised trial data by contacting the corresponding author.

## LIST OF ABBREVIATIONS

ART: antiretroviral therapy

UNAIDS: joint United Nations Programme on HIV/AIDS LMIC – low- and middle-income countries

VL: Viral Load

WHO: World Health Organization

## Author contributions

JD and NG conceived the study. YS, PKD, GH and CB contributed to study design. JD, KT, YS, PM, JN, ET, AM, NM, TM-G and NS contributed to study implementation. JN, PM and NS oversaw laboratory testing. LL created the allocation sequence and provided statistical support. JD and JAB conducted statistical analysis. JD wrote the first draft of the manuscript. All authors critically reviewed and edited the manuscript and consented to final publication.

## Acknowledgements

The authors would like to thank all participants in the study and acknowledge the work and support of staff at the Prince Cyril Zulu Clinic, eThekwini Municipality, CAPRISA and the National Health Laboratory Services at Addington and Inkosi Albert Luthuli Hospitals.

