## supplement for "Point-of-care HIV viral load testing in a community antiretroviral therapy delivery programme: a randomised controlled trial (PHILA)"

### Figure S1: Process maps of viral load testing in the PHILA trial

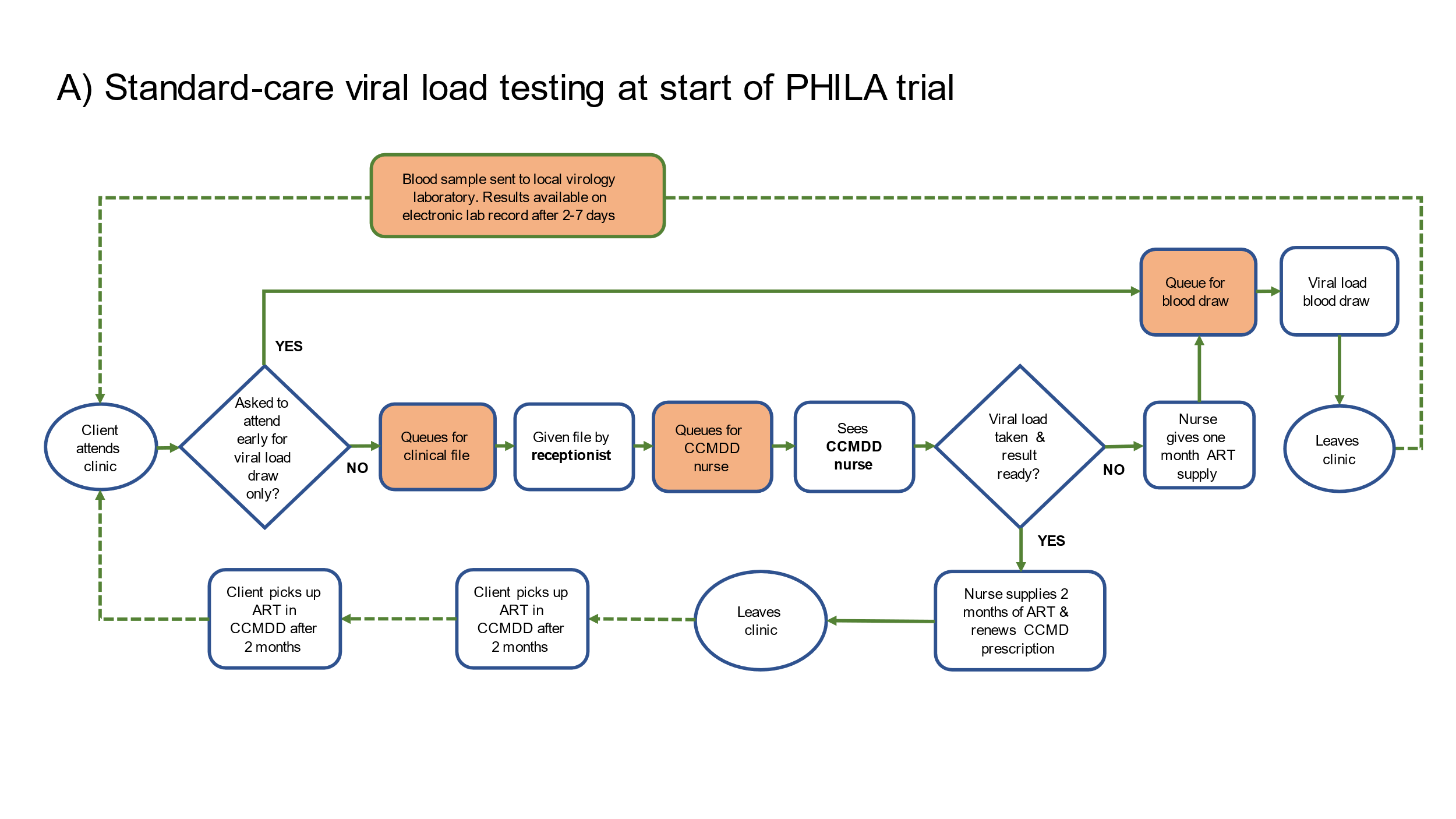

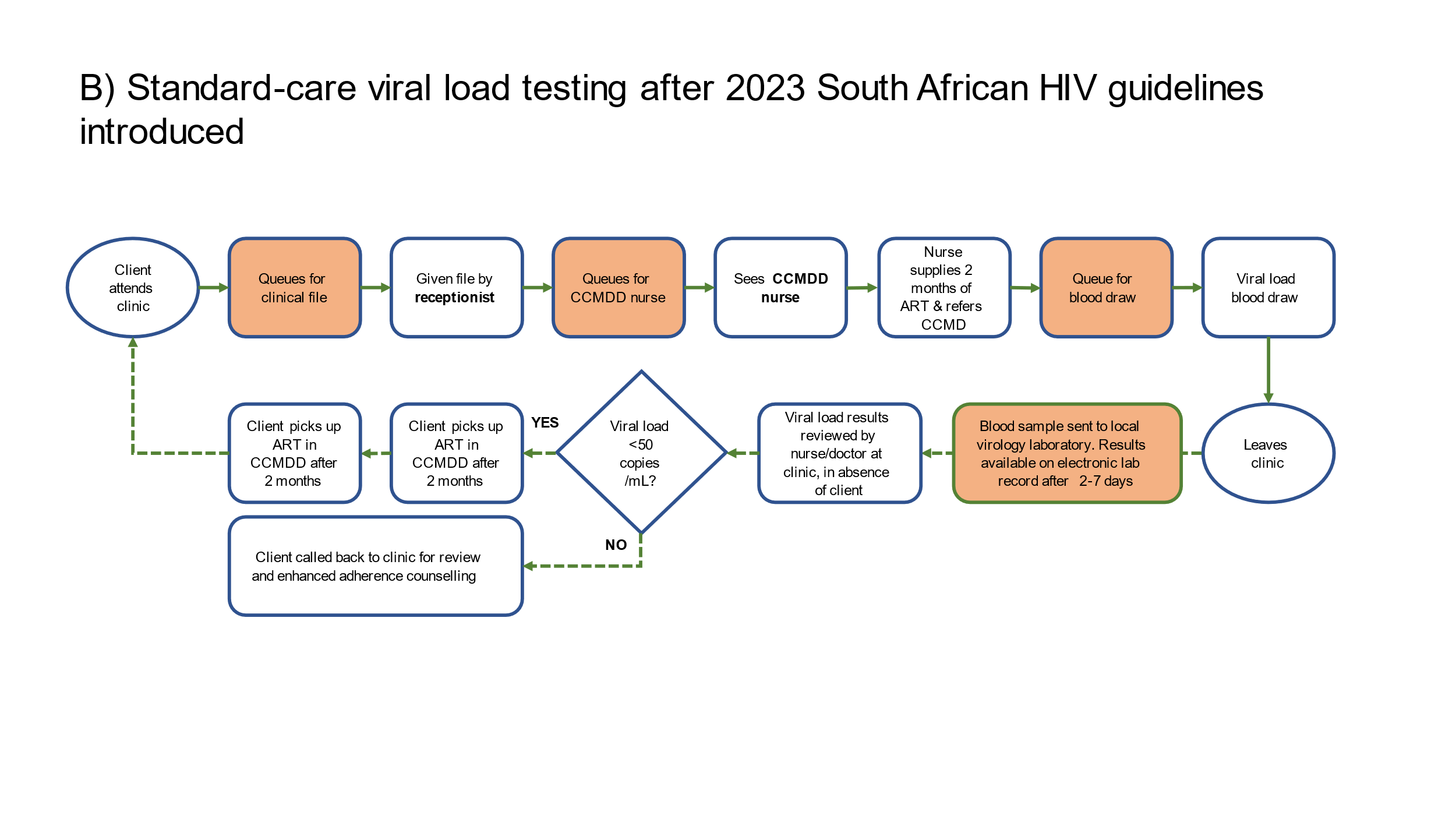

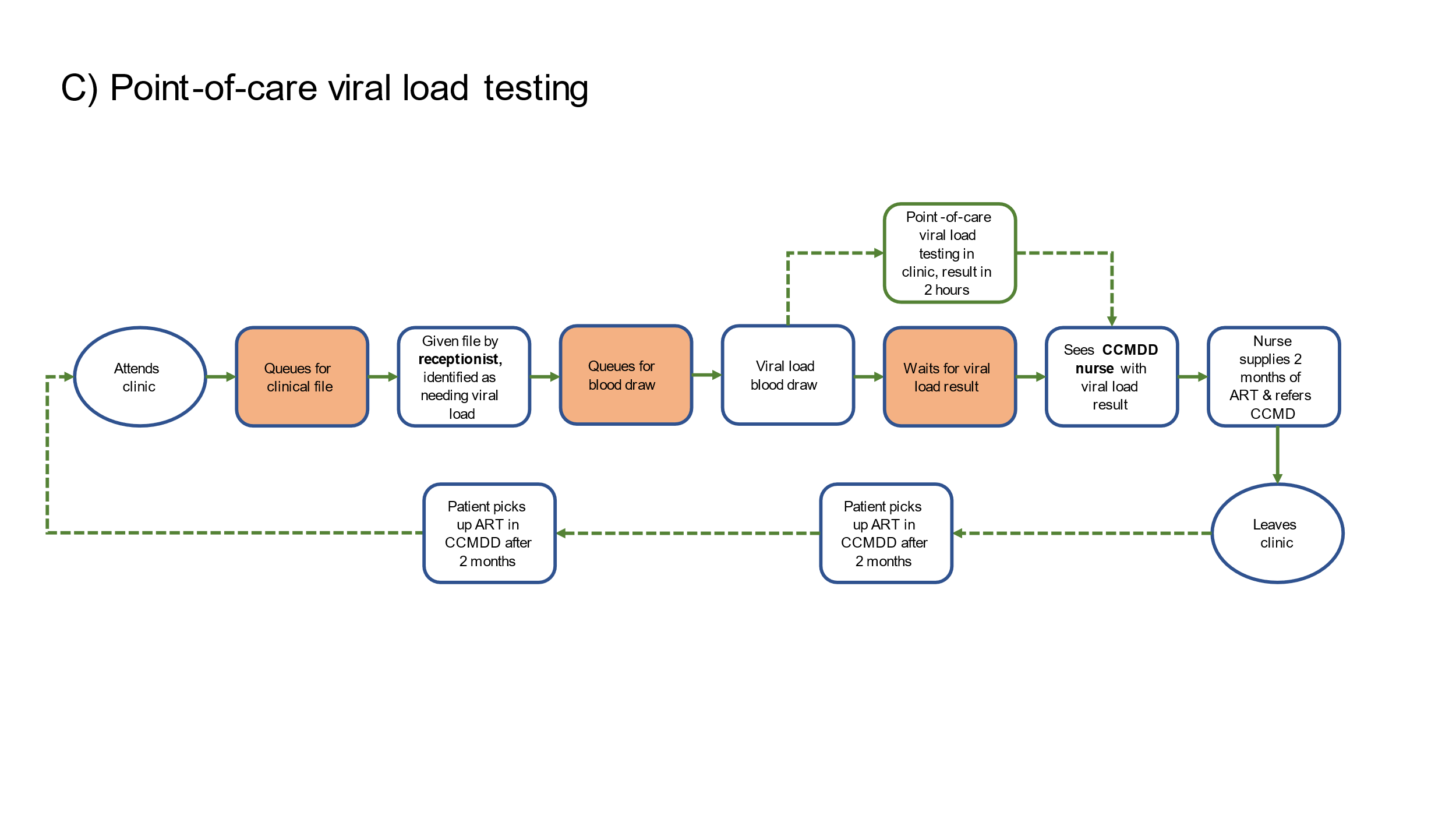

### Figure S2: Numbers of CCMDD prescriptions, and their cycle lengths, by month in the PHILA trial

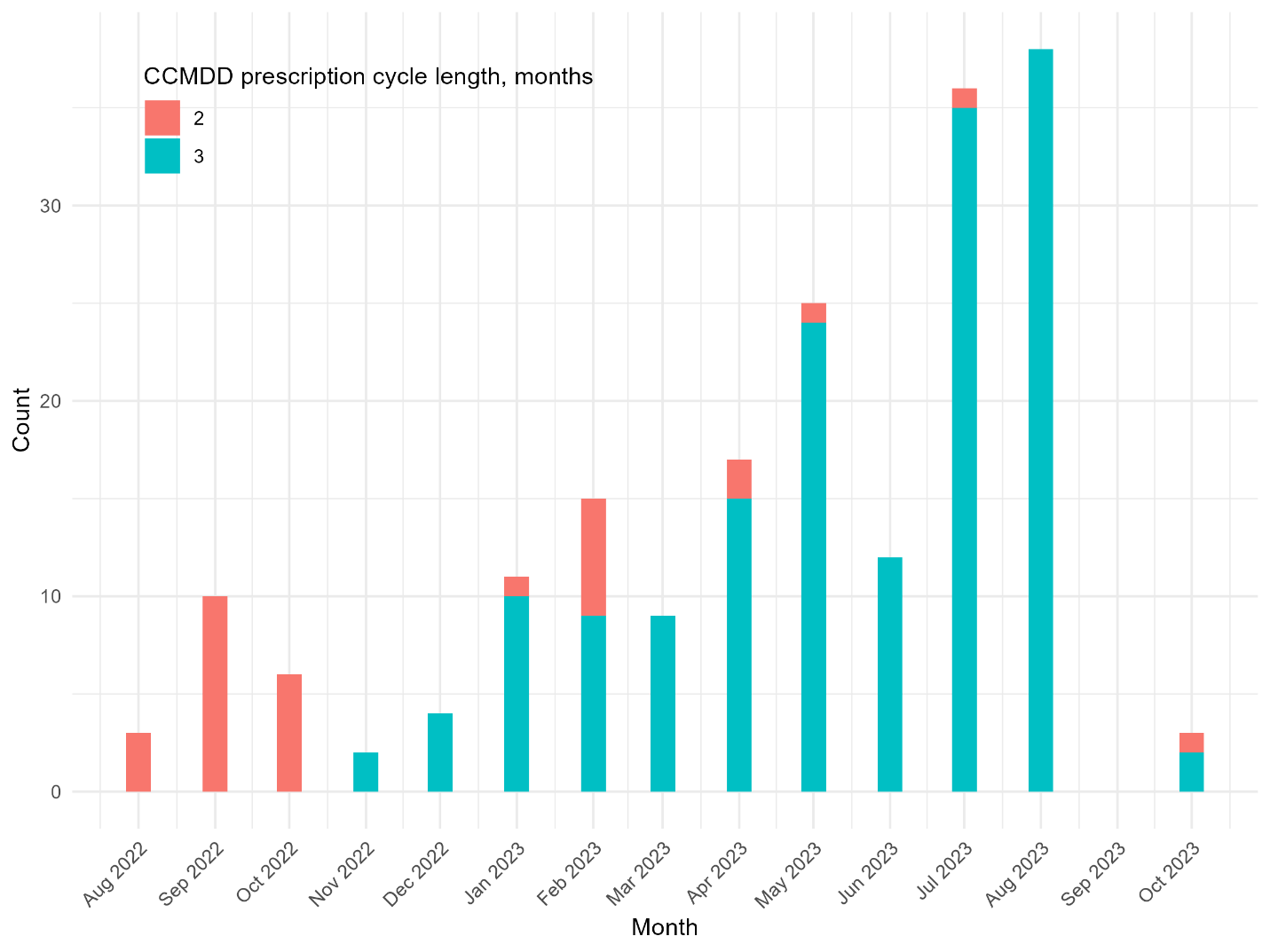

### Figure S3: Time to ART collection in CCMDD in the PHILA trial

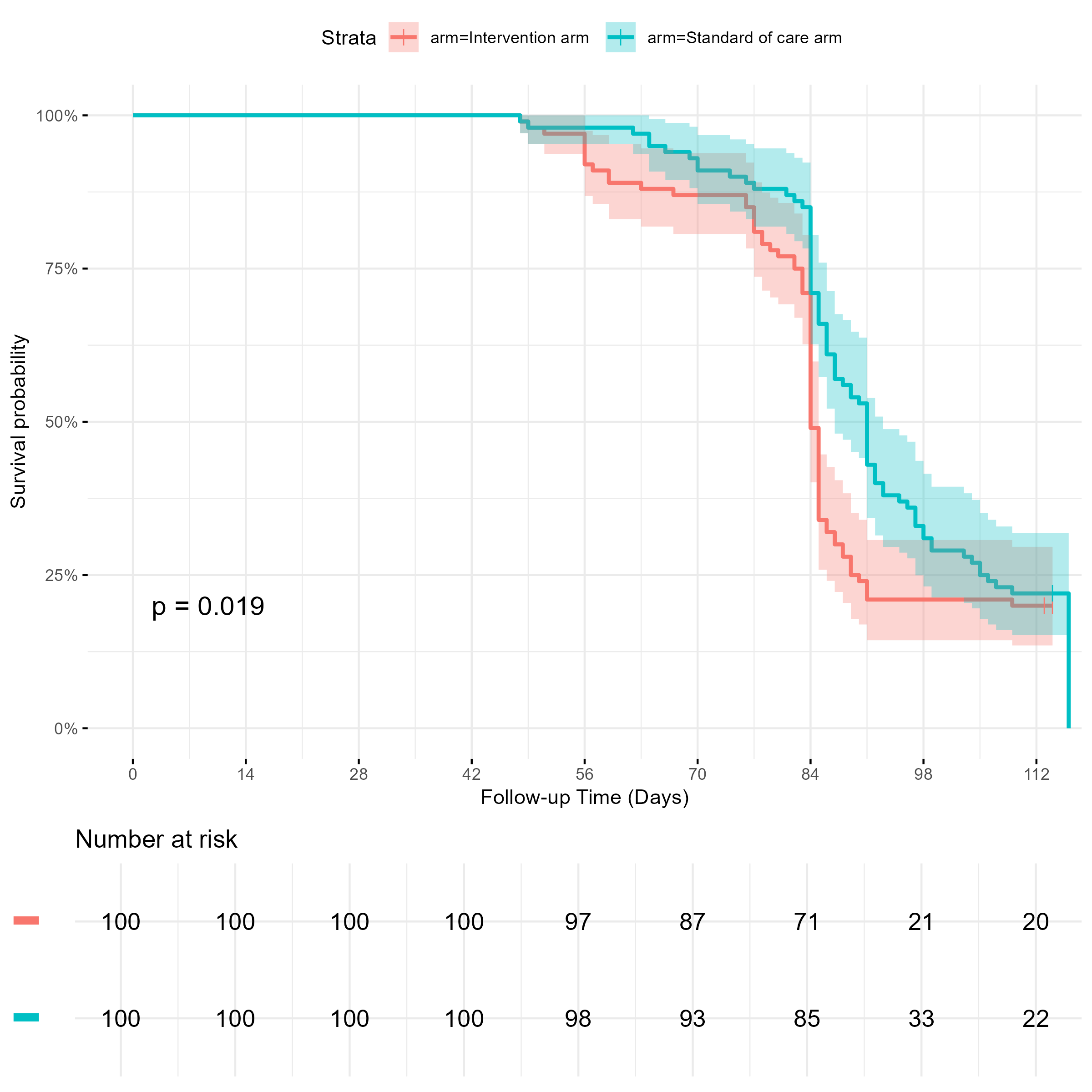

### Figure S4: Time to receipt of viral load results in the PHILA trial

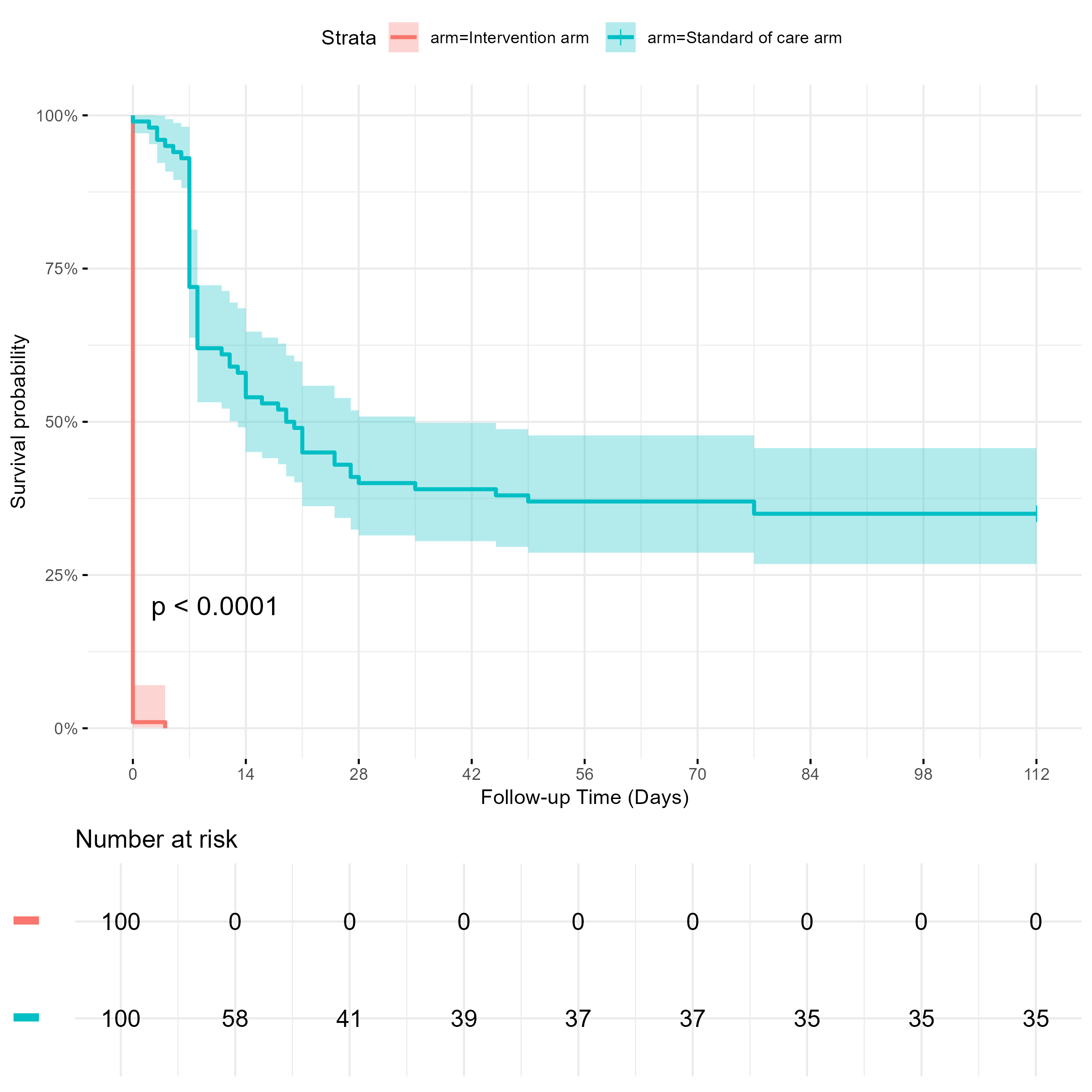

### Table S1: Reasons for not collecting antiretroviral therapy in the PHILA trial, n = 24

| **Description** | **N** |
| --- | --- |
| Participant not contacted | 13 |
| Participant travelling so couldn’t collect | 4 |
| Participant reported they had collected package, although no record of collection in SYNCH | 3 |
| Participant did not find antiretroviral therapy package at community pickup point | 1 |
| Participant died | 1 |
| Participant stated had enough ART so did not need to collect | 1 |
| Participant did not receive SMS reminder and so forgot to collect ART | 1 |

### Table S2: Characteristics of people receiving a renewed referral to CCMDD at the time of renewed referral in 108 facilities between 24^th^ May and 10^th^ September, n = 16,568

| **Variable** | **Levels** | **Overall**  **(n=16,568; column %)** | **Viral load at referral ≤50 copies/mL**  **(n=13,636; row %)** | **Viral load at referral >50 copies/mL**  **(n=2,932; row %)** |
| --- | --- | --- | --- | --- |
| **Demographics** | | | | |
| Age, years | Median (IQR) | 41 (35 to 48) | 41 (35 to 48) | 42 (37 to 49) |
| Gender | Female | 11,477 (69.3%) | 9,558 (83.3%) | 1,919 (16.7%) |
|  | Male | 5,091 (30.7%) | 4,078 (80.1%) | 1,013 (19.9%) |
| Region | eThekwini Metropolitan Municipality | 12,734 (76.9%) | 10,012 (78.6%) | 2,722 (21.4%) |
|  | uMgungundlovu District Municipality | 3,834 (23.1%) | 3,624 (94.5%) | 210 (5.5%) |
| **Clinical information** | | | | |
| Time since ART initiation, years | Median (IQR) | 7.9 (5.1 to 10.5) | 7.9 (5.1 to 10.3) | 8.0 (5.5 to 11.0) |
| Initiation CD4 count, cells/µL^1^ | Median (IQR) | 280 (160 to 440) | 290 (170 to 450) | 250 (140 to 370) |
| Initiation CD4 count category, cells/µL^1^ | <200 | 4,059 (32.0%) | 3,190 (78.6%) | 869 (21.4%) |
|  | 200-349 | 3,899 (30.7%) | 3,128 (80.2%) | 771 (19.8%) |
|  | 350-499 | 2,373 (18.7%) | 1,984 (83.6%) | 389 (16.4%) |
|  | >=500 | 2,360 (18.6%) | 2,067 (87.6%) | 293 (12.4%) |
|  | (Missing) | 3,877 | 3,267 | 610 |
| Current ART regimen at enrolment | TDF / XTC / DTG | 15,640 (94.4%) | 12,915 (82.6%) | 2,725 (17.4%) |
|  | TDF / XTC / EFV | 303 (1.8%) | 255 (84.2%) | 48 (15.8%) |
|  | Other | 625 (3.8%) | 466 (74.6%) | 159 (25.4%) |
| Time on current regimen, years | Median (IQR) | 2.5 (1.8 to 3.0) | 2.5 (1.7 to 3.0) | 2.5 (1.8 to 3.0) |
| Time since first CCMDD referral, years | Median (IQR) | 3.8 (1.8 to 5.6) | 3.8 (1.8 to 5.7) | 3.6 (1.8 to 5.5) |
| Time since latest CCMDD referral, days | Median (IQR) | 168 (168 to 172) | 168 (168 to 172) | 168 (168 to 172) |
| Time since previous viral load, days | Median (IQR) | 358 (335 to 378) | 360 (336 to 378) | 350 (286 to 379) |
| Previous viral load result, copies/mL | ≤50 | 14,985 (90.8%) | 12,618 (84.2%) | 2,367 (15.8%) |
|  | 51 - 999 | 1,443 (8.7%) | 918 (63.6%) | 525 (36.4%) |
|  | ≥1000 | 80 (0.5%) | 45 (56.3%) | 35 (48.3%) |
|  | (Missing) | 60 | 55 | 5 |

^1^ Closest to ART initiation, up to 180 days before and up to 30 days after ART initiation.
